# Trends in Homeschooling Rates Following Elimination of Nonmedical Exemptions to Childhood Immunizations, 2012-2020

**DOI:** 10.1101/2021.11.08.21266098

**Authors:** Kavin M. Patel, SarahAnn M. McFadden, Salini Mohanty, Caroline M. Joyce, Paul L. Delamater, Nicola P. Klein, Daniel A Salmon, Saad B. Omer, Alison M. Buttenheim

## Abstract

**Introduction:** In 2015 California passed Senate Bill No. 277 (SB 277) and became the first state in over 30 years to eliminate nonmedical exemptions to mandatory childhood immunizations for school entry. One concern that emerged was that the law created an incentive for parents to remove children from brick-and-mortar schools to bypass the immunization requirements.

**Objective:** To assess the impact of eliminating nonmedical exemptions to childhood immunizations on homeschooling rates.

**Design:** Pre-Post Intervention study. We calculated homeschooling rates as the number of K-8 students enrolled through each of California’s three homeschooling mechanisms (Independent Study Program, Private School Affidavit, and Private School Satellite Program) divided by all K-8 students enrolled in the same academic year. Data on homeschooling rates was obtained from the California Department of Education. We then conducted an interrupted time series analyses in which the outcome variable was percent of students enrolled in a homeschool program pre- and post-SB 277.

**Setting:** California homeschools

**Participants:** K-8 students enrolled through each of the state’s three homeschooling mechanisms (Independent Study Program, Private School Affidavit, and Private School Satellite Program)

**Intervention:** Passage of SB 277 which eliminated nonmedical exemptions to childhood immunizations for school entry

**Main Outcome:** K-8 homeschooling rates

**Results:** The homeschooling enrollment for K-8 students in California increased from 0.8% (35,122 students) during SY 2012-13 to 1.9% (86,574 students) during SY 2019-20; however, we found no significant increase in the percent of students enrolled in homeschooling programs in California following the implementation of SB 277 beyond the secular trend.

**Conclusions and Relevance:** Legislative action to limit nonmedical exemptions to compulsory vaccination for school entry is not associated with removal from classroom-based instruction in brick-and-mortar institutions.

**KEY POINTS:** *Question:* What is effect of California Senate Bill No. 277, which eliminated nonmedical exemptions to childhood immunizations for school entry, on the homeschooling rates?

*Finding:* We found no evidence that elimination of nonmedical exemptions to mandatory childhood immunizations for school entry was associated with an increase in homeschooling rates for K-8 in California.

*Meaning:* Legislative action to limit nonmedical exemptions to compulsory vaccination for school entry is not associated with removal from classroom-based instruction in brick-and-mortar institutions.

## INTRODUCTION

The so called Disneyland measles outbreak of 2015 followed a decade-long decline in childhood immunization coverage rates in California.^1^ The state has required childhood immunizations as a condition of school entry since 1977.^2^ However, there were two ways to bypass the mandate: (1) obtain a nonmedical exemption based on a personal and/or religious belief or (2) file for a medical exemption with the endorsement of a health care provider.^3-5^ A review of the Disneyland outbreak implicated personal belief exemptions as the reasons for under-or non-vaccination in two-thirds of measles cases.^6^ In response, state lawmakers passed Senate Bill No. 277 (SB 277)^7^ which eliminated nonmedical exemptions to childhood immunizations prior to the 2016-17 school year. SB 277 made California the first state in over 30 years to eliminate nonmedical exemptions^3^, thereby providing an opportunity to better understand both the intended and unintended consequences of such a policy change. School entry mandates and vaccine exemptions have been an active area of state policy and legislation,^8^ and many states have looked to California as an example when drafting similar legislation.

Following the implementation of SB 277, data from the California Department of Public Health^9,10^ showed that the law proved effective in increasing vaccine uptake with Kindergarten immunization rates increasing from 92.8% in 2015-16 to 95.1% in 2017-18.^11^ There was a concern, however, that the law created an incentive for parents who could no longer obtain a nonmedical exemption to either substitute a medical exemption^12^ or to homeschool their children in order to circumvent school entry immunization requirements (a new provision of SB277).^13^ In fact, the medical exemption rate for Kindergarteners almost tripled between 2015–2016 and 2017–2018^11,14^; when many of these were suspected of being fraudulent, the legislature passed Senate Bill No. 276 in 2019 to strengthen the requirements to obtain a medical exemption.^15^ While removal of unvaccinated children from congregate settings such as school and daycare may reduce the risk of highly transmissible vaccine-preventable diseases, it may also have consequences on children’s social and emotional development given differing opportunities for peer interaction.^16^ The purpose of this study was to evaluate the second potential mechanism to bypass vaccination by describing the changes in homeschooling rates before and after SB 277.

## METHODS

Parents choosing to homeschool in California may do so by one of several mechanisms; these include independent study programs (ISPs), private school satellite programs (PSPs), or by filing a private school affidavit (PSA) (Table 1).^17^ We describe below our approach to estimating enrollment for each homeschooling mechanism for school years (SY) 2012-13 to 2019-20.

**Table 1:** Overview of the Major Homeschooling Mechanisms in California

| Homeschooling Mechanism | Abbreviation | Type of School | Curriculum Support | Administrative Support | Fee | Secular | Reasons Parents May Choose Option |
| --- | --- | --- | --- | --- | --- | --- | --- |
| Traditional Independent Study Program | ISP | Public | Yes | No | No | Yes | To provide a structured education mirroring classroom-based instruction with a centralized curriculum composed of the recommended core subjects. |
| Charter School-Based Homeschool | ISP | Public | Yes (parents may choose to receive a stipend and purchase their own curriculum) | No | No | Yes | To provide a structured education usually adhering to a particular educational philosophy (i.e., Montessori and/or Waldorf) |
| Private School Affidavit | PSA | Private | No | No | No | No | To provide an individualized education at the parents' discretion. |
| Private School Satellite Program | PSP | Private | Variable | Yes | Yes | No | To provide a coordinated education that caters to a parent's and child's specific needs (frequently parochial). |

### Independent Study Program

An ISP is a program of the public school system in California. Compared to other homeschooling mechanisms, an ISP offers the greatest support to parents, by providing either a standardized curriculum and educational material or by providing funding so that parents may purchase their own curriculum.^18,19^ Parents may choose an ISP because their child has competing priorities (e.g., child actor, professional athlete, or child with health issues) or requires an adjusted curriculum (e.g., child requires specialized attention or qualifies for accelerated coursework) among other reasons.^18^ Students enrolled in an ISP are required to meet with a credentialed teacher regularly and take school- and state-administered tests.^19^ To identify ISP programs in California, we obtained a list of all past and current public schools from the California Department of Education (CDE).^20^ ISPs were identified as schools that offered any combination of grades inclusive of K-8 in at least one SY 2012-13 to 2019-20 and that were either “primarily virtual” (defined by the CDE as a school that provides virtual instruction but that may include some physical meetings between students and teachers) or “exclusively virtual” (defined by the CDE as a school where all instruction is virtual).^21^

### Private School Affidavit

Another mechanism for homeschooling in California is by filing a private school affidavit (PSA)^17,19,22^; this allows the family’s residence to become a standalone school with the parent acting as administrators and teachers. In this capacity, parents keep detailed records of attendance, coursework, and grades, and provide instruction in a variety of core subjects.^22^ PSAs receive no funding, curricular materials, or instructional support^23^ but afford a substantial degree of autonomy.^22^ While the CDE publishes data on private school enrollment, these data are censored for schools with fewer than six students. To obtain the censored data, we filed a California Public Records Act request with the CDE for a de-identified listing of all schools with five or fewer enrolled students. We created two estimates of homeschooling enrollment via PSA: The ‘low estimate’ included schools that had an enrollment of only one student; whereas the ‘high estimate’ included schools that had enrollment of 5 or fewer students.

### Private School Satellite Program

A Private School Satellite Program (PSP) is a private school that has filed an affidavit and whose main function is to support home-based instruction.^24,25^ PSPs may require parents to serve as teachers and/or administrators, and provide administrative, curricular, and community support (including opportunities for students to socialize and engage in enrichment activities) in exchange for a membership fee.^17,24,26^ Some PSPs are denominational.^25^ California maintains a database of all private schools in state^27^; however, the database does not distinguish between a brick-and-mortar private school and a PSP. To identify PSPs, we compiled a list from PSP directories maintained on the websites of three homeschooling networks (California Homeschool Network^24^, A2Z Homeschooling^23^, Homeschooling Concierge^25^.) To confirm that the listed schools were PSPs, we individually reviewed each school’s website and/or contacted the school administrators via phone or email. Of note, there were 15 PSPs that did not exist in the CDE database. An additional 9 PSPs were brick-and-mortar schools with a PSP offshoot; these were excluded as it is unclear what proportion of the total enrollment was accounted for by the PSP. To address uncertainty in PSP classification, we created two estimates of PSP enrollment: The “low estimate” included only schools that were confirmed as PSPs via website, phone call or email communication with a school official, whereas the “high estimate” also included schools that could not be confirmed as PSPs.

### Total Enrollment

The total number of students enrolled in grades K-8 in California was calculated by combining the count data from three separate databases: the public-school database (available on the CDE public school website^20^), the private school database with five or fewer students (available from the CDE through a California Public Records Act request), and the private school database with greater than five students (available on the CDE private school website^27^). The CDE databases also contained a category of “ungraded elementary” which represented students who did not fit into a particular grade level; these were included in the numerator for ISP/PSA/PSP counts and in the denominator for total enrollment.

### Data Analysis

We calculated homeschooling rates as the number of children enrolled through each of California’s three homeschooling mechanisms divided by all students enrolled in school in the same academic year. We calculated rates for each school year 2012-13 through 2019-20 for the entire K-8 population, as well as grade-specific and homeschooling mechanism-specific (ISP, PSA, PSP) rates. Rates were calculated for both low and high estimates of PSP and PSA enrollment as described above.

To evaluate the impact of the implementation of SB277 on the percent of children homeschooled in California, we conducted six interrupted time series analyses in which the outcome variable was percent of children enrolled in a homeschool program. Time periods (SB277) were defined as pre-SB277 (academic years 2012-13 – 2015-16) and post-SB277 (academic years 2016-17 – 2019-2020; dummy coded 0 and 1, with 0 being pre-SB277), given the 2016 implementation date of the law. Other variables included in the model were academic year (year) as well as a composite variable of homeschooling mechanism (ISP, PSA, PSP) and grade level (K-8), which was used as a random intercept to account for previous trends in homeschool rates.

The outcomes for the six analyses were as follows: low and high estimates for all children enrolled in all homeschool programs, low and high estimates for all children enrolled in kindergarten for all homeschool programs, K-8 children enrolled in ISPs, and kindergartners enrolled in ISPs. Restriction of the sample to K-8 enrollment in ISPs alone evaluated the hypothesis that parents seeking to homeschool their children merely to bypass vaccination would be more likely to choose the option with the greatest degree of curricular or financial support. Restriction to kindergarten enrollment only evaluated the hypothesis that any effects of the elimination of nonmedical exemptions would be seen at the main entry point into schooling. Students already enrolled with a nonmedical exemption prior to SB277 were grandfathered in subsequent grades.^28^. Inferential analyses were conducted in Stata^©^ (16.1, StataCorp LLC, College Station, TX).

## RESULTS

### Total Homeschooling Enrollment

Total homeschooling enrollment for K-8 students in California increased from 35,122 students or 0.8% of all enrolled K-8 students in the state in SY 2012-13 to 86,574 students or 1.9% during SY 2019-20 for the low estimate (42,379 students or 0.9% to 97,316 students or 2.1% for the high estimate).

### Homeschooling Enrollment by Grade Level

The increase in homeschooling in California was greatest for the lower versus the upper grade levels; for example, kindergarten homeschooling enrollment had increased from 2,068 students or 0.4% of the total in SY 2012-13 to 10,553 students or 1.9% in SY 2019-20 (2,231 students or 0.4% to 10,969 students or 1.9% for the high estimate), while Grade 8 homeschool enrollment rate increased from 5,146 students or 1.0% in SY 2012-13 to 10,485 students or 2.0% in SY 2019-20 (5,263 students or 1.0% to 10,666 students or 2.0% for the high estimate).

### Homeschooling Enrollment by Mechanism

The mechanisms responsible for the majority of homeschooling during the year SB 277 was enacted (SY 2015-16) included the following from most common to least common: ISP – 20,149 students or 45% (20,149 students or 38% for high estimate); PSA – 19,333 students or 44% (27,989 students or 53% for high estimate); and PSP – 4,733 students or 11% (4,935 students or 9% for high estimate).

### Homeschooling Enrollment Pre- and Post-SB 277

We found no significant increase in the percent of children enrolled in homeschooling programs in California following the implementation of SB277 in 2016 (*Tables 2 and 3*). However, when comparing pre-SB277 implementation to post-SB277 implementation, we did find small decreases in the percent change of students in all grade levels enrolled in ISPs (β = -0.0065, CI = -0.0088 – -0.0043; *Table 2a*) and specifically for kindergarteners enrolled in ISPs (β = -0.008 CI = -0.0097 – -0.0057; *Table 2b*).

**Table 2a.**
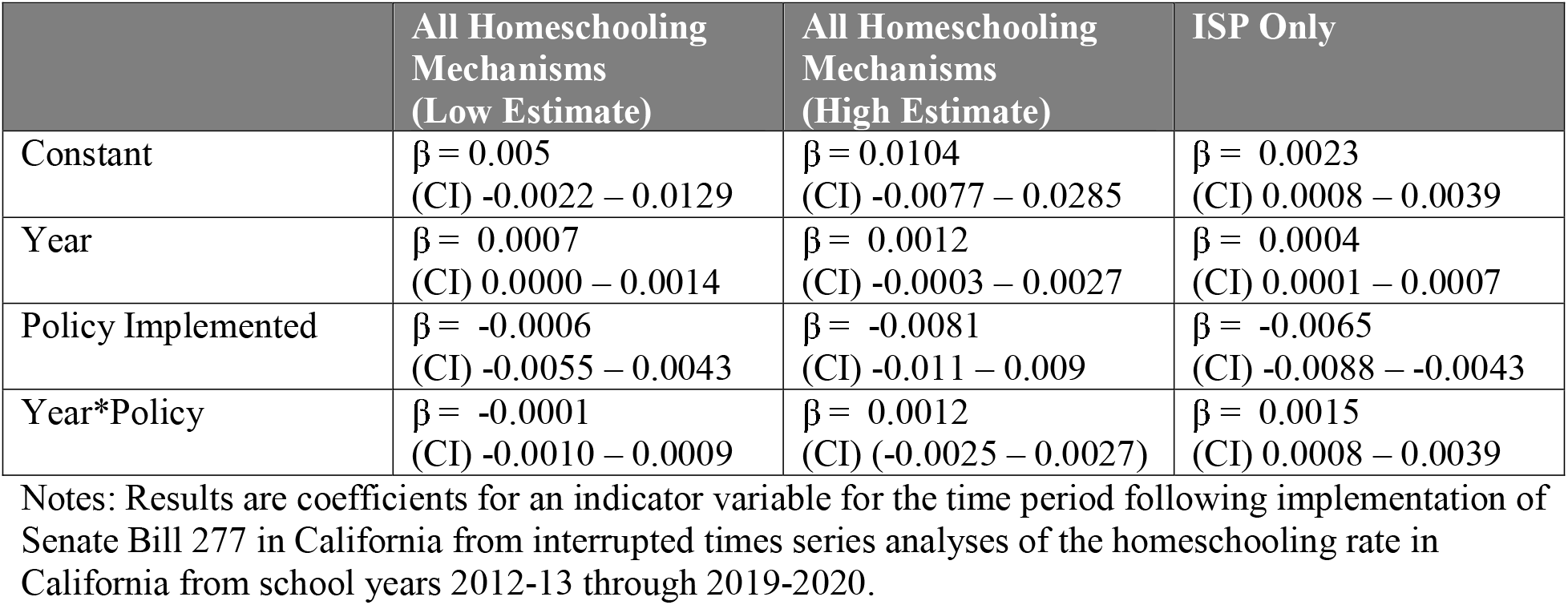
Effect of Elimination of Nonmedical Exemptions in California via SB277 in 2016 on ISP/PSA/PSP enrollment for K-8 students

|  | All Homeschooling Mechanisms (Low Estimate) | All Homeschooling Mechanisms (High Estimate) | ISP Only |
| --- | --- | --- | --- |
| Constant | $\beta = 0.005$<br>(CI) -0.0022 – 0.0129 | $\beta = 0.0104$<br>(CI) -0.0077 – 0.0285 | $\beta = 0.0023$<br>(CI) 0.0008 – 0.0039 |
| Year | $\beta = 0.0007$<br>(CI) 0.0000 – 0.0014 | $\beta = 0.0012$<br>(CI) -0.0003 – 0.0027 | $\beta = 0.0004$<br>(CI) 0.0001 – 0.0007 |
| Policy Implemented | $\beta = -0.0006$<br>(CI) -0.0055 – 0.0043 | $\beta = -0.0081$<br>(CI) -0.011 – 0.009 | $\beta = -0.0065$<br>(CI) -0.0088 – -0.0043 |
| Year*Policy | $\beta = -0.0001$<br>(CI) -0.0010 – 0.0009 | $\beta = 0.0012$<br>(CI) (-0.0025 – 0.0027) | $\beta = 0.0015$<br>(CI) 0.0008 – 0.0039 |

Notes: Results are coefficients for an indicator variable for the time period following implementation of Senate Bill 277 in California from interrupted times series analyses of the homeschooling rate in California from school years 2012-13 through 2019-2020.

**Table 2b.** Effect of Elimination of Nonmedical Exemptions in California via SB277 in 2016 on ISP/PSA/PSP enrollment for kindergarten students

|  | All Homeschooling Mechanisms (Low Estimate) | All Homeschooling Mechanisms (High Estimate) | ISP Only |
| --- | --- | --- | --- |
| Constant | $\beta = 0.0010$<br>(CI) -0.0028 – 0.0048 | $\beta = 0.0011$<br>(CI) -0.0026 – 0.0049 | $\beta = 0.0012$<br>(CI) 0.0004 – 0.0019 |
| Year | $\beta = 0.0002$<br>(CI) -0.0008 – 0.0013 | $\beta = 0.0002$<br>(CI) -0.0008 – 0.0012 | $\beta = 0.0005$<br>(CI) 0.0002 – 0.0008 |
| Policy Implemented | $\beta = -0.0039$<br>(CI) -0.0113 – 0.0034 | $\beta = -0.0042$<br>(CI) -0.0115 – 0.0032 | $\beta = -0.008$<br>(CI) -0.0097 – -0.0057 |
| Year*Policy | $\beta = 0.0009$<br>(CI) -0.0006 – 0.0024 | $\beta = 0.0010$<br>(CI) -0.0027 – 0.0048 | $\beta = 0.0020$<br>(CI) 0.0016 – 0.0024 |

Notes: Results are coefficients for an indicator variable for the time period following implementation of Senate Bill 277 in California from interrupted times series analyses of the homeschooling rate in California from school years 2012-13 through 2019-2020.

## DISCUSSION

This is the first study, to our knowledge, that estimates homeschooling rates in California. We found no evidence that SB 277’s elimination of nonmedical exemptions to mandatory childhood immunizations for school entry prior to the 2016-17 school year was associated with an increase in homeschooling rates for K-8 in California. This suggests that SB 277 had minimal unintended consequences related to homeschooling.^3^ This is reassuring as it may indicate that legislative action to limit exemptions to compulsory vaccination for school entry is not associated with removal from classroom-based instruction in brick-and-mortar institutions nor restricted opportunities for peer interactions.

Our results are consistent with multiple explanations of parents’ response to SB 277. One possibility is that vaccine-hesitant or vaccine-refusing parents who would have pursued a nonmedical exemption if available decided that the effort of homeschooling outweighed their opposition to vaccination. Alternatively, parents who would have pursued a nonmedical exemption, if available, may have instead pursued a medical exemption – an outcome consistent with the observed tripling of the medical exemption rate between 2015–2016 (pre-SB 277) and 2017–2018 (post-SB 277), documented by our team.^14^ Another drastic mechanism for avoiding SB 277’s restrictions – moving out of the state – is difficult to document as a result of SB 277. Finally, parents may have avoided homeschooling as a solution to SB 277 simply because schools did not enforce SB 277 consistently. In previous work, we found evidence to suggest that the law was variably interpreted, implemented, and enforced across school districts given vague regulatory language.^11,29^ One study found that the number of students “overdue” for vaccination more than quadrupled^3^, suggesting that some parents were being allowed to enroll their children in brick-and-mortar schools despite being under-or unvaccinated.

It is important to note that other legislative, regulatory, and surveillance responses to increasing nonmedical exemptions in California may have created pressure to homeschool prior to SB 277. For example, Assembly Bill 2109 (passed in 2014, implemented in 2015) tightened the criteria for obtaining a nonmedical exemption.^3^ In 2015, state and local health departments in California additionally began an effort to ensure proper application of the state’s conditional school entrance criteria for students not up to date with vaccinations.^3^ If this was the case, increases in homeschooling rates may have occurred over several years rather than immediately following SB 277’s implementation, creating the possibility of a type 2 error in our analysis. However, we find this explanation unlikely given that prior to SB 277, children who were homeschooled via either public or private homeschooling options were subject to the same school entry immunization mandates and had the same nonmedical exemptions options available to them.

We note one important limitation to our study: while we are confident that ISP and PSA data are complete and accurate, PSP data are subject to reporting biases inherent in crowdsourced listings on websites. Additional possible sources of error include PSPs that were either not registered with the CDE (and had no publicly available data); or PSPs that were affiliated with brick-and-mortar private schools (and had merged data). Given the relatively small contribution of PSPs to overall homeschooling rates (approximately 10 percent), we do not believe the uncertainties around PSP enrollment had a significant impact on the analysis.

## CONCLUSION

A review of the homeschooling trends in California before and after SB 277 suggests that the elimination of nonmedical exemptions to mandatory childhood immunizations for school entry was not associated with an increase in homeschooling rates. This conclusion is predicated upon the continued ability of vaccine refusing parents to enroll children in brick-and-mortar schools either by substituting with a medical exemption or by residing in a local school district with poor implementation of the law. States looking to eliminate nonmedical exemptions to childhood immunizations can learn from California’s example to better understand some of the unintended consequences that may result upon elimination of nonmedical exemptions to childhood immunizations; they may then use this insight to craft legislation capable of realizing the largest gains in vaccination rates with minimal unintended consequences.

## Data Availability

All data produced are available online at: https://www.cde.ca.gov/ds/

## Abbreviations

SB 277: Senate Bill No. 277
ISP: Independent Study Program
PSA: Private School Affidavit
PSP: Private School Satellite Program
CDE: California Dept. of Education

## DECLARATIONS

### Conflict of Interest Disclosures

NPK reports research support from Pfizer, Merck, GlaxoSmithKline, Sanofi Pasteur, and Protein Science (now Sanofi Pasteur) for unrelated studies.

### Funding/Support

Supported by grant R01AI125405 from the National Institutes of Health. The National Institutes of Health had no role in the design and conduct of the study, management, analysis, and interpretation of the data, or preparation of the final manuscript. Funded by the National Institutes of Health (NIH).

### Authors’ Contributions

Dr. Patel performed the data collection and drafted the initial manuscript. Dr. Buttenheim conceptualized the study, provided critical oversight, and reviewed the manuscript for important intellectual content. Dr. Mohanty and Ms. Joyce participated in data collection and preliminary analyses. Dr. McFadden performed the data analysis and reviewed the manuscript for important intellectual content. Drs. Mohanty, Delamater, Klein and Salmon critically reviewed the manuscript for important intellectual content. Dr. Omer is the principal investigator and helped to conceptualize the study and reviewed the manuscript for important intellectual content. All authors approved the final manuscript as submitted.

## TABLES & FIGURES

**Figure 1:**
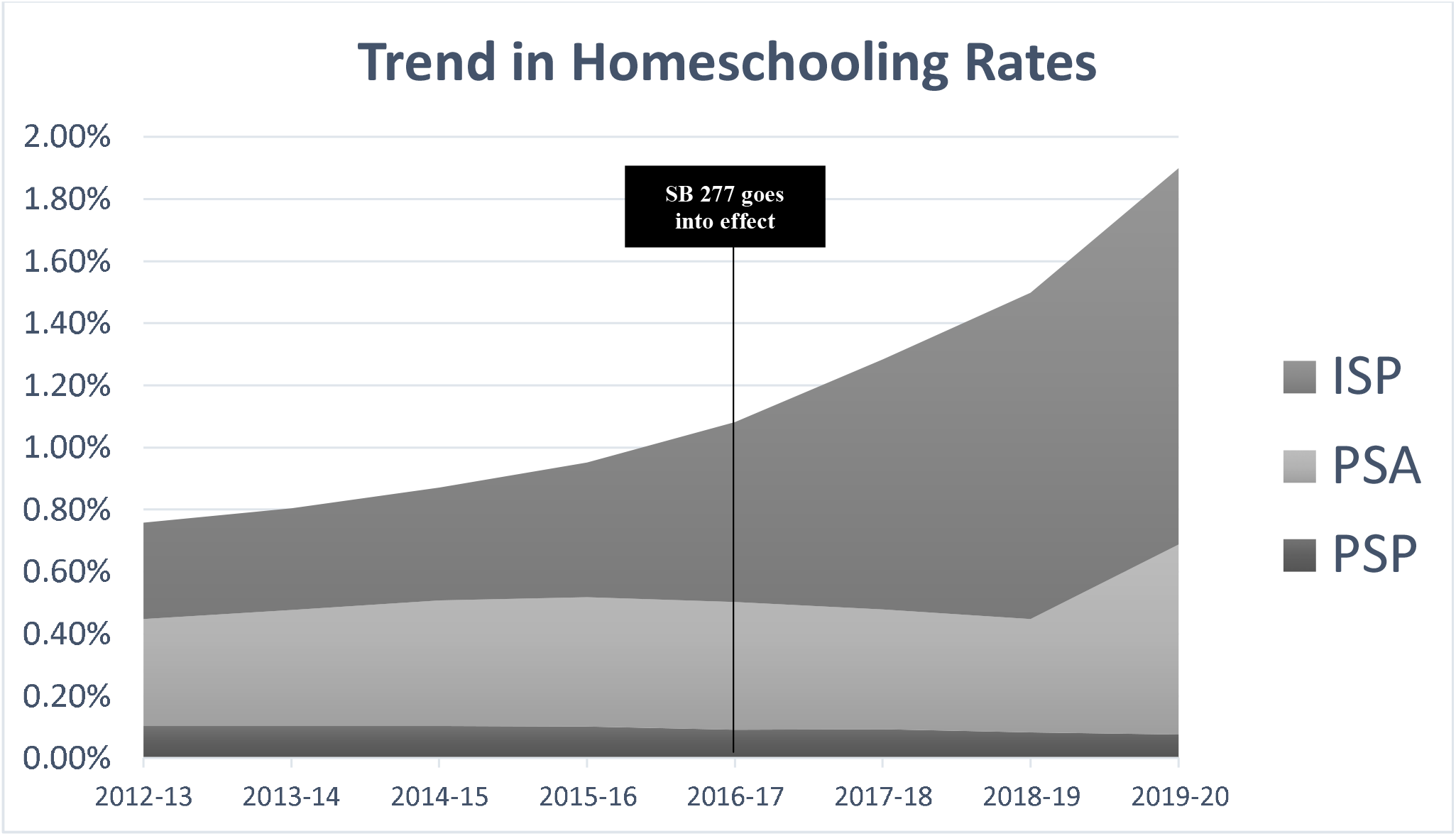
Trends in Homeschooling Rates in California for School Years 2012-13 to 2019-20.

**Figure 2:**
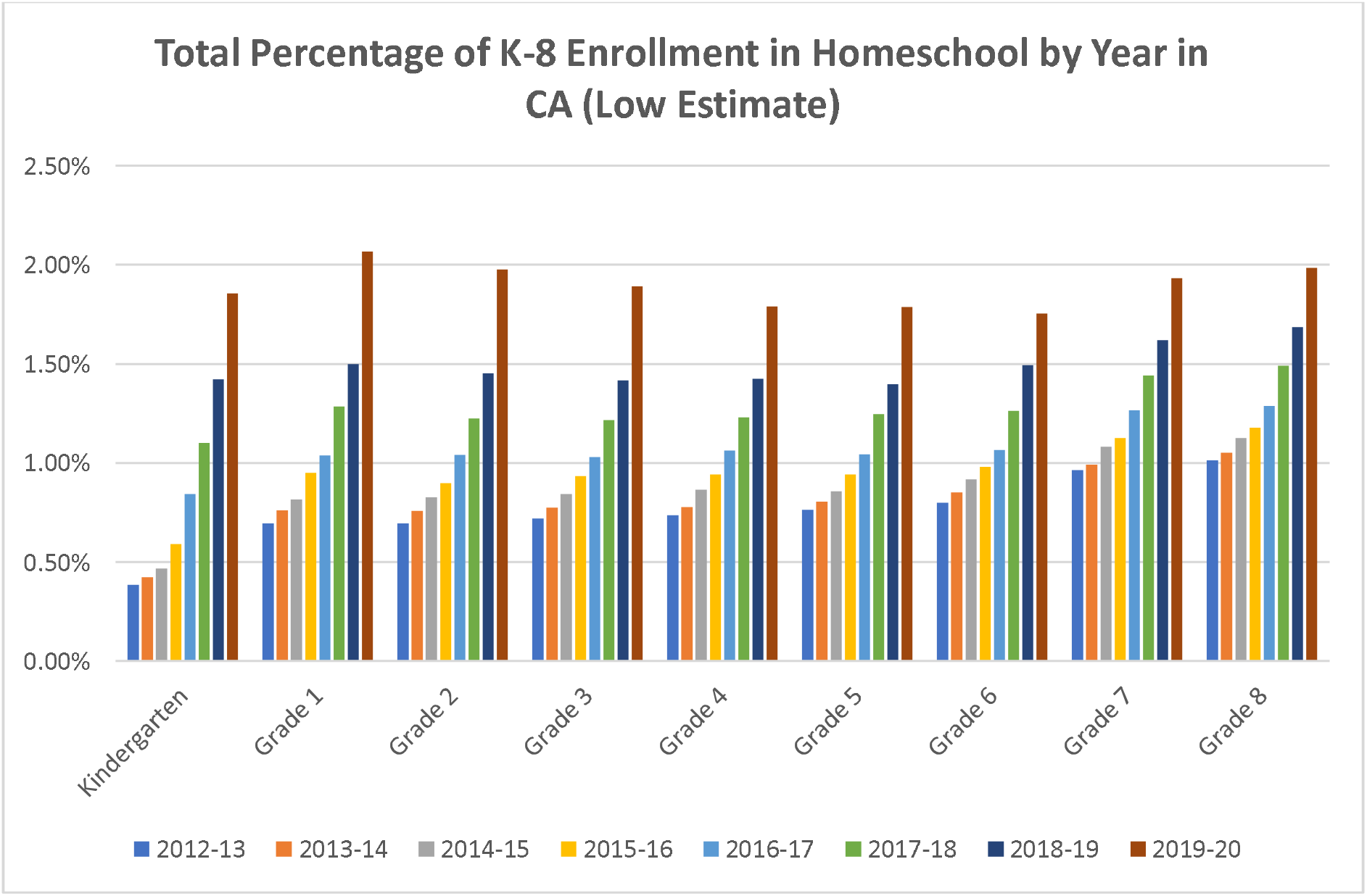

## Notes

### Author Declarations

California Department of Education

